# A needle in a haystack: metagenomic DNA sequencing to quantify *Mycobacterium tuberculosis* DNA and diagnose tuberculosis

**DOI:** 10.1101/2022.04.15.22273912

**Authors:** Adrienne Chang, Omary Mzava, Liz-Audrey Djomnang Kounatse, Joan Lenz, Philip Burnham, Peter Kaplinsky, Alfred Andama, John Connelly, Christine M. Bachman, Adithya Cattamanchi, Amy Steadman, Iwijn De Vlaminck

## Abstract

**Background:** Tuberculosis (TB) remains a significant cause of mortality worldwide. Metagenomic next-generation sequencing has the potential to reveal biomarkers of active disease, identify coinfection, and improve detection for sputum-scarce or culture-negative cases.

**Methods:** We conducted a large-scale comparative study of 427 plasma, urine, and oral swab samples from 334 individuals from TB endemic and non-endemic regions to evaluate the utility of a shotgun metagenomic DNA sequencing assay for tuberculosis diagnosis.

**Findings:** We found that the choice of a negative, non-TB control cohort had a strong impact on the measured performance of the diagnostic test: the use of a control patient cohort from a nonendemic region led to a test with nearly 100% specificity and sensitivity, whereas controls from TB endemic regions exhibited a high background of nontuberculous mycobacterial DNA, limiting the diagnostic performance of the test. Using mathematical modeling and quantitative comparisons to matched qPCR data, we found that the burden of *Mycobacterium tuberculosis* DNA constitutes a very small fraction (0.04 or less) of the total abundance of DNA originating from mycobacteria in samples from TB endemic regions.

**Interpretation:** Our findings suggest that the utility of a metagenomic sequencing assay for tuberculosis diagnostics is limited by the low burden of *M. tuberculosis* in extrapulmonary sites and an overwhelming biological background of nontuberculous mycobacterial DNA.

**Funding:** This work was supported by the National Institutes of Health, the Rainin Foundation, the National Science Foundation, and the Bill and Melinda Gates Foundation.

## Introduction

Despite the introduction of the World Health Organization’s (WHO) End TB Strategy in 2015, tuberculosis (TB) remains one of the top global causes of death due to a single infectious pathogen. In 2021, the WHO reported the first rise in deaths due to tuberculosis in over a decade.^1^ Recent advances in nucleic acid amplification tests, including the WHO-recommended Xpert® MTB/RIF Ultra, have produced rapid tests with high positive predictive value^2^. However, these assays are unable to replace the use of culture for bacteriological confirmation due to low negative predictive value or implementation barriers present in low- and middle-income countries that account for 98% of reported TB cases^1^. Though sputum culture has long been a gold standard for TB diagnostics, the time to positive detection can be affected by a number of variables, including sputum volume, population characteristics (e.g. age, HIV coinfection), and method of specimen handling^3,4^. Variations due to these factors can lead to discrepancies between quantitative and semiquantitative diagnostic assays. Thus, there is an unmet need for robust, point-of-care tuberculosis diagnostics.

Metagenomics DNA sequencing has the capacity to detect all potential infectious pathogens in a clinical sample using an unbiased approach. Numerous studies have demonstrated that DNA from *Mycobacterium tuberculosis*, the causative agent of tuberculosis, can be detected in plasma, urine, and oral fluids^5–7^. However, its diagnostic performance in distinguishing positive sputum culture from negative sputum culture samples has fallen short of the performance standards set by the WHO (98% specificity, 80% sensitivity)^8^. We explored the utility of a metagenomic sequencing assay for tuberculosis across three biofluids and four geographic regions and found that diagnostic performance is highly influenced by the geographic origin of the control cohort (**Fig 1**). Using simulation and modeling, we found that the diagnostic performance is correlated with the abundance of *M. tuberculosis* DNA relative to the background of DNA from nontuberculous mycobacteria. The background of DNA originating from nontuberculous mycobacteria is low in samples from TB non-endemic regions but overwhelms and obscures the *M. tuberculosis* signal in samples from TB endemic regions. Our study provides insight into the burden and properties of *M. tuberculosis* in different biofluids and can inform the development of molecular tests that achieve the requisite standards for sensitivity and specificity.

**Figure 1.**
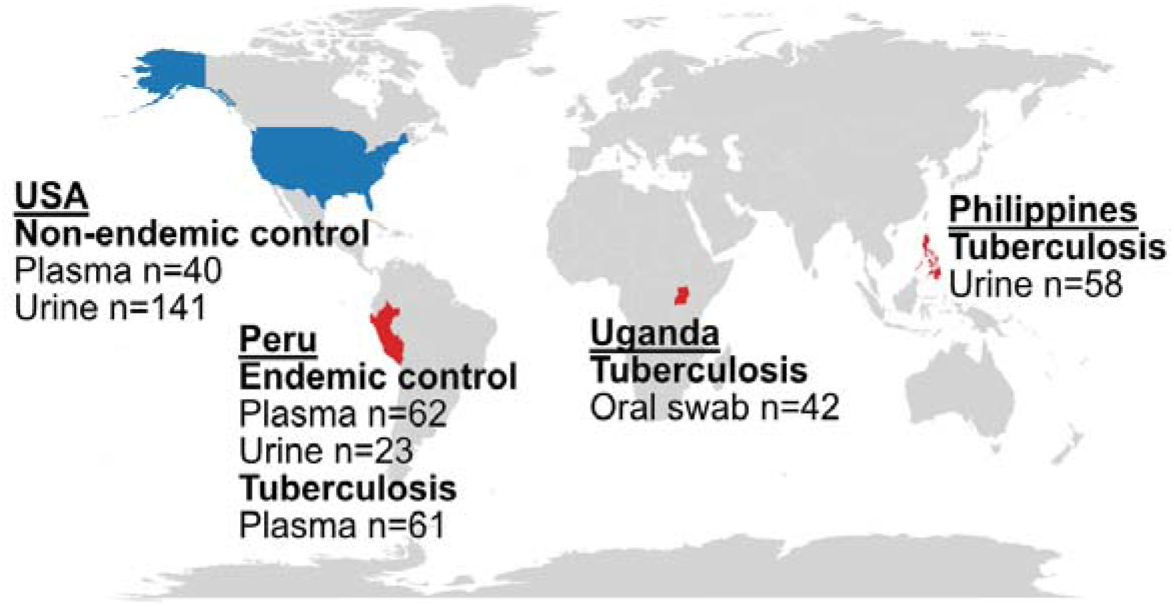
Geographic distribution of samples included in this study. High tuberculosis burden countries are shaded in red.

## Results

### *Biophysical properties of* M. tuberculosis *DNA in urine, plasma, and oral swabs*

Microbial cell-free DNA (cfDNA) in urine and plasma has been shown to be ultrashort, with an average fragment length of less than 100 bp (**Fig 2A**)^9,10^. Compared to these biofluids, relatively few studies have examined the properties and diagnostic potential of microbial DNA from oral swabs. Oral swabs samples are an attractive new avenue for metagenomic DNA assays because they can be obtained noninvasively, are more cost effective than obtaining, storing, and shipping blood samples, and provide a high yield of DNA^11^. We found that in contrast to the microbial fragment length profiles of urinary and plasma cfDNA, DNA obtained from oral swabs is longer and likely genomic in origin: the fragmentation pattern closely mirrors that of human chromosomal DNA which arises from the tagmentation step in the library preparation protocol (**Fig 2B**).

**Figure 2.**
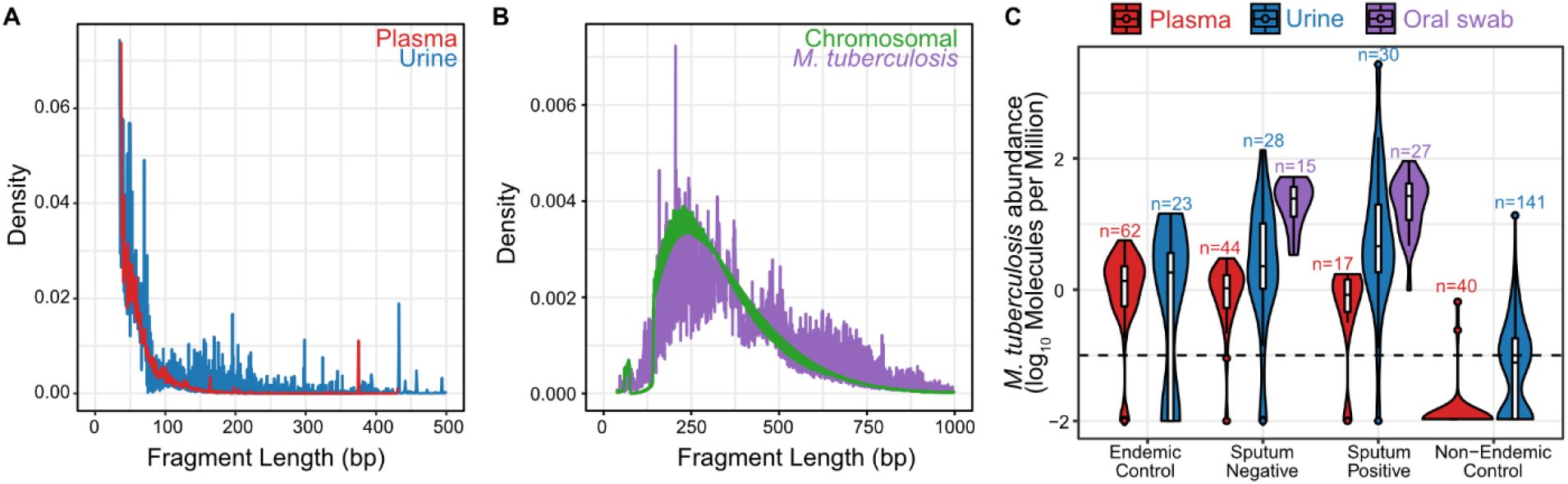
**A** The fragment length distributions of *M. tuberculosis* DNA in plasma (red) and urine (blue). **B** The fragment length distribution of *M. tuberculosis* DNA (purple) and host chromosomal DNA (green) in oral swabs are similar, suggesting that the microbial DNA is genomic and the fragmentation profile is not an intrinsic property but rather the consequence of sample preparation steps. **C** The abundance of *M. tuberculosis* DNA across all cohorts and biofluids. The majority of non-endemic samples have little to no detectable *M. tuberculosis* DNA, while the abundance of *M. tuberculosis* DNA in endemic and tuberculosis samples increases from plasma to oral swab. Dashed line indicates a limit of detection cutoff of 0.1 MPM.

We set out to quantify the abundance of *M. tuberculosis* DNA detected in 161 samples obtained from patients presenting with symptoms of respiratory illness at tuberculosis clinics in the Philippines and Uganda (tuberculosis cohort), approximately half of which were sputum positive for tuberculosis (**Table 1**). We found that the abundance of *M. tuberculosis* DNA increases from plasma, to urine, to oral swabs (**Fig 2C**). Within the tuberculosis cohort, there was no difference in *M. tuberculosis* DNA abundance between sputum positive and sputum negative tuberculosis samples. Setting a limit of detection of 0.1 molecule per million reads (MPM), we find that within the tuberculosis cohort we detect *M. tuberculosis* DNA in 100% of the oral swab samples (42/42), 95% of the urine samples (55/58), and 93% of the plasma samples (57/61). The median abundance of *M. tuberculosis* DNA in the tuberculosis cohort was 0.512, 2.29, and 25.5 MPM for plasma, urine, and oral swabs, respectively. Further, we found an average of 27.9-fold more (adjusted p-value = 3.3×10^−16^, Wilcoxon test) *M. tuberculosis* molecules in the oral swab samples than in plasma samples.

**Table 1.** Overview of datasets included in this study. All data was generated for this study unless otherwise indicated.

| Cohort | Origin | Disease | Biofluid | Patients | Samples |
| --- | --- | --- | --- | --- | --- |
| Endemic | Peru | Environmental enteropathy | Plasma | 62 | 62 |
|  |  |  | Urine | 23 | 23 |
| Non-endemic | USA | Kidney transplant<sup>12</sup> | Urine | 82 | 141 |
|  |  | Lung transplant<sup>13,14</sup> | Plasma | 6 | 40 |
| Tuberculosis | Philippines<sup>10</sup> | Sputum positive | Urine | 30 | 30 |
|  |  | Sputum negative | Urine | 28 | 28 |
|  | Peru | Sputum positive | Plasma | 17 | 17 |
|  |  | Sputum negative | Plasma | 44 | 44 |
|  | Uganda | Sputum positive | Oral swab | 27 | 27 |
|  |  | Sputum negative | Oral swab | 15 | 15 |

As a point of comparison, we quantified the abundance of *M. tuberculosis* in plasma and urine samples from two additional cohorts: 1) 141 urine samples and 40 plasma samples from patients living in the United States who received kidney or lung transplants, respectively^12–14^(non-endemic cohort); and, 2) 62 plasma samples and 23 urine samples from pediatric patients with suspected environmental enteropathy in Peru, a TB endemic region (endemic cohort). We found no *M. tuberculosis* DNA signatures in 38/40 plasma non-endemic samples and 89/141 urine non-endemic samples. In contrast, *M. tuberculosis* DNA was detected in 57/62 plasma samples and 16/23 urine samples from the endemic cohort. The median abundance of *M. tuberculosis* DNA in the endemic cohort was 1.26 and 1.85 MPM in plasma and urine samples, respectively. The higher abundance of *M. tuberculosis* DNA in the endemic cohort relative to the non-endemic cohort was expected given that geography, ethnicity, and other population-based factors have previously been shown to influence the microbiome composition among healthy individuals^15^, and that the presence of an infectious organism does not necessarily equate to a disease state^16^. Given that the abundance of *M. tuberculosis* DNA across all biofluids in the endemic cohort was over 3.7-fold more than the non-endemic cohort (adjusted p-value = 3.4×10^−14^, Wilcoxon test), but still significantly less than the tuberculosis cohort (12.15-fold less across all biofluids; adjusted p-value = 7 × 10^−4^, Wilcox test).

### *Diagnostic potential of* M. tuberculosis *DNA is influenced by choice of controls*

We evaluated the diagnostic performance of *M. tuberculosis* DNA in oral swabs, plasma, and urine. We found poor separation between sputum positive and sputum negative individuals across all biofluid types (area under the curve [AUC] = 0.6, 0.6, and 0.5 for plasma, urine, and oral swabs, respectively; **Fig 3A**). We found a modest increase in performance when comparing the tuberculosis and endemic cohorts (AUC = 0.62 and 0.64 for urine and plasma, respectively; **Fig 3B**). In contrast, we found almost perfect separation for the tuberculosis and non-endemic cohorts (AUC = 0.93 and 0.97 for urine and plasma, respectively; **Fig 3C**). Moreover, we found that the diagnostic performance was not influenced by the choice of metagenomic classifier, reference database, or removal of confounding reads (see **Table S1**-**S2**, and Supplemental Information).

**Figure 3.**
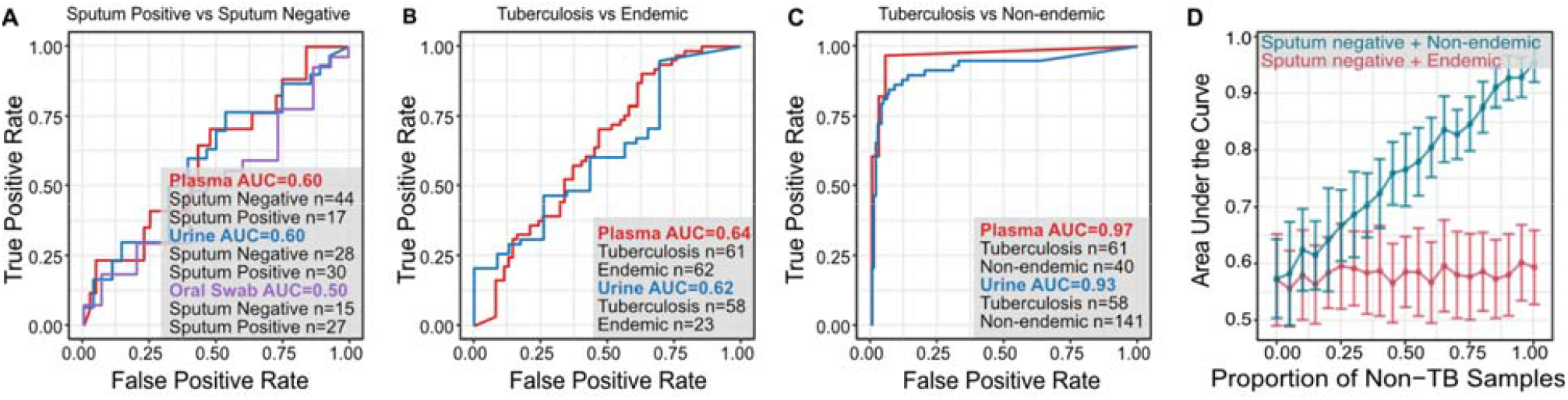
The performance of the metagenomic assay in discriminating **A** sputum positive versus sputum negative samples, **B** tuberculosis versus endemic cohorts, and **C** tuberculosis versus non-endemic cohorts (AUC = area under the curve). **D** Evaluating the effects of incorporating non-endemic samples (blue) and endemic samples (red) on the diagnostic performance via simulation show that the inclusion of non-endemic samples skews the assay’s sensitivity.

To explore the effects of a biological background on the diagnostic performance, we randomly selected combinations of 15 urine samples composed of sputum positive and a known mixture of sputum negative and non-endemic controls. We performed 50 sampling rounds and found a positive correlation between the proportion of non-endemic samples and the diagnostic performance, with a mean AUC of 0.57 when the negative control consisted of only sputum negative samples and a mean AUC of 0.95 when the negative control was composed entirely of non-endemic samples (**Fig 3D**). However, we saw no correlation when we performed the same analysis using a mixture of sputum negative tuberculosis and endemic controls: the area under the curve remained relatively constant, fluctuating between 0.56 and 0.60 across all negative control mixtures of sputum negative and endemic samples. This observation highlights a crucial but often overlooked criterium for metagenomic diagnostic test development: the choice of control. Differences in diagnostic performance are highly influenced by the geographic origin of the samples. This is supported by the performance of published studies assaying nucleic acids in blood or urine for tuberculosis diagnosis: sputum culture positive and sputum culture negative samples are nearly indistinguishable unless the control cohort include samples from individuals living in TB non-endemic regions (**Table S3**).

### Poor diagnostic performance can be attributed to a background of biological nontuberculous mycobacteria

We hypothesized that the poor diagnostic performance could be attributed to geographic factors: individuals living in TB endemic regions are exposed to nontuberculous mycobacteria, such as *M. avium, M. abscessus*, and *M. kansasii*^17^. This is further supported by observations that the abundance of *M. tuberculosis* DNA is positively correlated with age (**Fig S1**). This exposure results in a nontuberculous mycobacteria background that is indistinguishable from a true disease signal due to the low abundance of *M. tuberculosis* and the high sequence similarity between *Mycobacterium* genomes, which can exceed 99% nucleotide similarity^18^.

To determine whether the poor diagnostic performance could be attributed to a biological nontuberculous mycobacteria background, we simulated datasets by digitally spiking in *M. tuberculosis* reads into datasets generated for non-endemic, endemic, and tuberculosis urine samples. Across a range of 0 to 1000 spiked-in *M. tuberculosis* reads, we obtained nearly perfect classification of the synthetic reads in non-endemic samples (0.987 ± 0.0210; **Fig 4A**). However, the endemic and tuberculosis samples exhibited logarithmic relationships between the number of spiked-in reads and precision. When we evaluated precision as a function of the relative coverage of spiked-in *M. tuberculosis* reads to all *Mycobacterium* reads in a sample, we found a strong correlation between the signal-to-noise ratio and the classification precision (**Fig 4B**). Non-endemic samples had little to no background DNA from *Mycobacterium* species and exhibited high precision, while endemic and tuberculosis samples had a higher background of nontuberculous mycobacteria that reduced the classification precision. To further test our hypothesis that background nontuberculous mycobacterial DNA drives poor classification precision, we removed all *Mycobacterium* reads prior to spiking in *M. tuberculosis, M. bovis*, or *M. avium* reads and found perfect classification across all samples, regardless of the number of reads simulated (**Tables S4-S6**).

**Figure 4.**
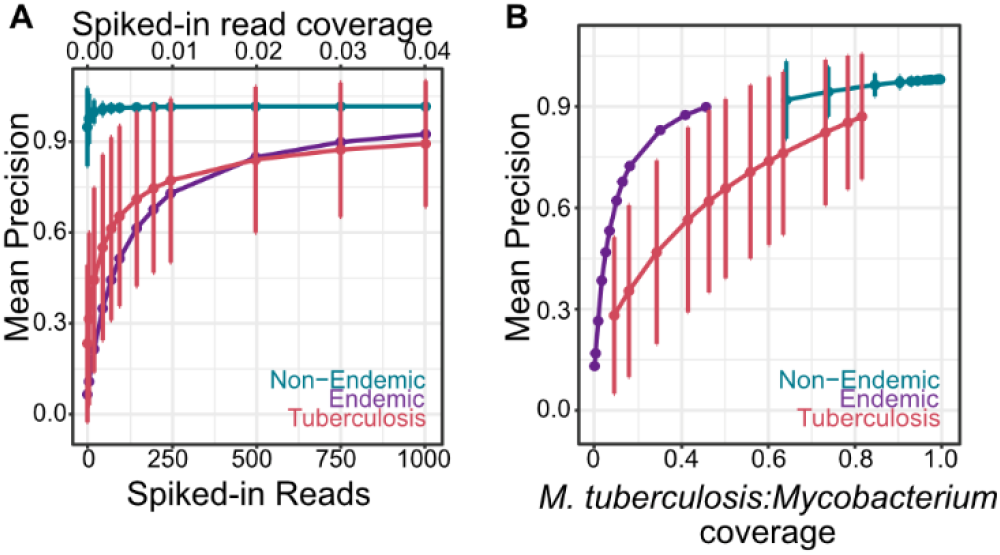
**A** Classification precision of synthetic *M. tuberculosis* reads spiked into non-endemic urine samples (n=29, 5 replicates per sample), endemic urine samples (n=23, 5 replicates per sample), and tuberculosis urine samples (n=66, 5 replicates per sample). **B** The classification precision is correlated with the relative coverage of *M. tuberculosis* to total *Mycobacterium* in the sample.

The availability of sputum PCR results for the oral swab samples provided further opportunity to evaluate the *M. tuberculosis* signal relative to the nontuberculous mycobacterial background. The Xpert® MTB/RIF Ultra assay (Cepheid, Sunnyvale, USA) targets three segments of the *M. tuberculosis* complex, two of which are the IS6110 and IS1081 insertion sequences. *M. tuberculosis* isolates contain between 0-25 copies of IS6110^19^, whereas the IS1081 is present in all *M. tuberculosis* complex species at a stable number of 5-7 repeats per genome^20^. We evaluated the presence of IS1081 detected by metagenomic sequencing because the range in copy numbers across different *M. tuberculosis* complex species is narrower. IS1081 was detected by metagenomic sequencing in 5 of 27 sputum positive oral swab samples and was not detected in any of the 15 sputum negative oral swab samples, as expected. Because IS1081 is unique to *M. tuberculosis*, we were able to obtain a lower bound of the relative abundance of *M. tuberculosis* versus *Mycobacterium* by comparing the per-base sequence coverage of the IS1081 gene segment relative to the *M. tuberculosis* genome. Using the minimum copy number for the IS1081 gene (five copies per genome), we found that the coverage of nontuberculous mycobacteria relative to IS1081 was 210.397 (range of relative coverage: 23-394). Given that IS1081 was not detected in 22 of the 27 sputum positive oral swab samples and was not detected in any of the 15 sputum negative oral swab samples, this range represents a lower bound. Thus, the burden of *M*. tuberculosis DNA represents 4.4% or less of the total abundance of *Mycobacterium* DNA, indicating a significant background of nontuberculous mycobacteria. Using species-specific insertion sequences revealed that the background of *Mycobacterium* originates from a number of species, all of which are both ubiquitous environmental mycobacteria and implicated in nontuberculous mycobacterial lung infections (*M. branderi, M. smegmatis, M. avium, M. celatum, M. gordonae, M. xenopi, M. fortuitum, M. ulcerans;* **Table S7**)^17,21^. Further validation is required to determine if these species are co-infectious and influence disease outcome.

## Discussion

We show that the utility of a metagenomic sequencing assay for tuberculosis diagnostics is dependent on the geographic origin of control samples and limited by the low abundance of *M. tuberculosis* in extrapulmonary sites. Such an assay is sensitive to the detection of nontuberculous mycobacteria that arises from a lifelong exposure to species from the *Mycobacterium* genus and contributes to the microbiome composition of samples originating from TB endemic regions. Our findings demonstrate that the influence of geography on the microbiome directly impacts the diagnostic performance: the inclusion of non-endemic samples as controls invariably results in a near perfect test while poor diagnostic separation is obtained when endemic samples are employed as controls. Mathematical modeling demonstrates that the diagnostic potential is correlated with the abundance of *M. tuberculosis* DNA relative to the background of nontuberculous mycobacterial DNA. Quantitative comparisons to matched qPCR reveals that nontuberculous mycobacterial DNA is 23-fold or more abundant than the abundance *M. tuberculosis* DNA in the samples investigated here. The overwhelming biological background of *Mycobacterium* in samples of interest, in combination with the low abundance of *M. tuberculosis* in extrapulmonary sites, presents a major barrier for the implementation of an unbiased metagenomic DNA sequencing assay for tuberculosis diagnostics.

Detection of tuberculosis using a metagenomic sequencing assay for tuberculosis diagnostics is thus akin to looking for a needle in a haystack: *M. tuberculosis* DNA constituted less than 4.4% of the total abundance of *Mycobacterium* in samples from TB endemic regions included in this study. Our work suggests that the median abundance of *M. tuberculosis* is lower than 0.06 and 0.42 copies/mL in blood and urine, respectively, and lower than 284 genome copies/µg of DNA collected by oral swab, an estimate that is in agreement with previous reports quantifying the abundance of *M. tuberculosis* DNA through sequence-specific amplification^7,22^. Improvements to a metagenomic sequencing assay for tuberculosis diagnostics could be made by increasing the volume of input biofluid^23^, choosing a sample preparation workflow with improved DNA extraction and short read amplification^10,22^, or enriching for *M. tuberculosis-*specific sequences using ultrashort PCR amplicons^7,24^. These approaches would minimize noise from nontuberculous mycobacteria and increase the sequencing budget allocated to *M. tuberculosis*. Additionally, further exploration of the nontuberculous mycobacteria fraction may reveal patterns in disease outcome and provide new insights in the development of a robust geographic control. Together, our results reveal challenges and opportunities for the development of a DNA-based diagnostic tests for tuberculosis and provides a comprehensive characterization of *M. tuberculosis* in extrapulmonary sites that can inform the development of molecular tests.

## Methods

### Study cohorts

A total of 427 datasets from plasma, urine, and oral swab samples were analyzed, 188 of which were generated for this study (Table 2). Endemic plasma and urine samples were collected from patients seeking treatment for environmental enteropathy in Peru. The study was approved by the Johns Hopkins Bloomberg School of Public Health Institutional Review Board (protocol 00002185) and the Cornell University Institutional Review Board (protocol 1612006853). Tuberculosis plasma samples were collected from patients presenting with respiratory symptoms from tuberculosis clinics in Peru. The study was approved by the Foundation for Innovative New Diagnostics’ Clinical Trials Review Committee and the Cornell Institutional Review Board (protocol 1612006851). Oral swab samples were collected from individuals who presented with symptoms of respiratory illness at outpatient clinics in Uganda. The study was approved by the Makerere University School of Medicine Research and Ethics Committee (protocol 2017-020).

Additional datasets collected in the scope of previous studies were included as follows. Non-endemic urine samples were collected from kidney transplant patients who received care at New York Presbyterian Hospital-Weill Cornell Medical Center. The study was approved by the Weill Cornell Medicine Institutional Review Board (protocols 9402002786, 1207012730, 0710009490)^12^. Non-endemic plasma samples were collected from lung transplant patients who received care at Stanford University Hospital. The study was approved by the Stanford University Institutional Review Board (protocol 17666)^13,14^. Tuberculosis urine samples from patients seeking treatment for tuberculosis in the Philippines were collected through a study partly funded by the Department of Science and Technology - Philippine Council for Health Research and Development (DOST-PCHRD). This study was approved by the University of the Philippines Manila Research Ethics Board (protocol UPMREB 2018-252-01)^10^. All patients provided written informed consent.

### Sample collection

Urine samples were collected via the conventional clean-catch midstream method. For the endemic cohort, approximately 50 mL of urine was centrifuged at 3,000 *x g* on the same day for 30 minutes and the supernatant was stored in 1 mL aliquots at -80°C. For the tuberculosis cohort, approximately 10 mL of urine was mixed with 2 mL Streck Cell-Free DNA Urine Preserve (Streck, Cat #230604) and centrifuged at 3,000 *x g* for 30 minutes at ambient temperature within 30 minutes of specimen collection. The supernatant was similarly stored in 1 mL aliquots at -80°C.

Peripheral blood samples were collected in EDTA. Plasma was separated by centrifugation at 1,600 *x g* for 10 minutes followed by centrifugation at 16,000 *x g* for 10 minutes. Plasma was stored in 1 mL aliquots at -80°C.

Oral swab samples were collected by swabbing the tongue for 15 seconds with rotation using a Copan regular flocked swab with molded breakpoint at 30 mm (Copan, Cat #520CS01). The swab head was then broken off into a collection tube containing 1X TE buffer, vortexed for 30 seconds, and stored at -80°C.

### DNA isolation and library preparation

DNA was extracted from plasma and urine using the QIAamp Circulating Nucleic Acid Kit according to the manufacturer’s instructions (Qiagen, Cat #55114). Sequencing libraries were prepared using a single-stranded library preparation as described in Burnham et al^9^. DNA was extracted from oral swab samples using the QIAamp UCP Pathogen Kit according to the manufacturer’s instructions (Qiagen, Cat #50214) and libraries were prepared using the Nextera XT DNA Library Prep Kit (Illumina, Cat #FC-131-1024). All libraries were characterized using the AATI Fragment Analyzer before pooling and sequencing on the Illumina NextSeq 500 platform (paired-end, 2×75 bp).

### Fragment length distribution and quantification of M. tuberculosis

Low-quality bases and Illumina-specific sequences were trimmed (Trimmomatic-0.32, LEADING:25 TRAILING:25 SLIDINGWINDOW:4:30 MINLEN:15)^25^. Reads were aligned (BWA-mem^26^) to the human reference (UCSC hg19). Reads that did not align to the human genome reference were extracted and aligned to the bacteriophage phiX174.

To obtain the fragment length distribution for *M. tuberculosis* DNA, unaligned reads were extracted again and aligned to the circularized *M. tuberculosis H37Rv* genome (edited from NC_000962.3). Reads aligning with a minimum mapping quality of 30 were extracted, and the lengths computed as the absolute difference between the start and end coordinates. The fragment length density profile was computed using the *hist* function from the R Graphics Package.

To quantify the abundance of *M. tuberculosis*, reads that did not align to the human and phiX references were extracted and aligned to a curated list of bacterial and viral reference genomes using kallisto (--pseudobam)^27,28^. Reads aligning to the 16S/23S ribosomal RNA region were removed. Microbial abundance was quantified as Molecules per Million reads (MPM):

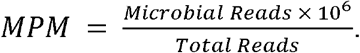

### Quantification of insertion sequence and genome coverage

Reads that did not align to the human and phiX references were extracted and aligned (BW-mem^26^) to the circularized *M. tuberculosis H37Rv* genome (edited from NC_000962.3), to the IS6110 and IS1081 sequences retrieved from the GenBank repository for *M. tuberculosis* H37Rv (NC_000962.3), and to additional nontuberculous mycobacteria-specific insertion sequences (**Table S7**). Reads aligning with a minimum mapping quality of 30 were extracted, and the lengths computed as the absolute difference between the start and end coordinates. The coverage was calculated using the following equation:

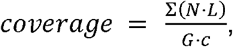

where *N* is the number of reads of length *L, G* is the sequence length, and *c* is the sequence copy number.

### ROC analysis

Receiver operating characteristic analyses were performed using the *roc* function the R package pROC^29^. For each biofluid, the abundance of *M. tuberculosis* DNA (in MPM) was compared between sputum positive tuberculosis and sputum negative tuberculosis samples, between tuberculosis and endemic cohorts, and between tuberculosis and non-endemic cohorts.

To evaluate the effects of using non-endemic and endemic samples on the diagnostic performance for tuberculosis, reads that aligned to *M. tuberculosis* were extracted from each sample. Sputum positive tuberculosis samples were compared to a known mixture of sputum negative tuberculosis samples and either non-endemic or endemic samples. For each of 50 sampling rounds, 15 samples were randomly chosen for ROC analysis.

### Simulating microbial DNA reads

Paired-end reads were simulated according to microbial cfDNA fragment length distribution from the *M. tuberculosis H37Rv* (NC_000962.3), *M. bovis BCG str. Pasteur 1173P2* genome (NC_008769.1), and *M. avium subsp. Hominissuis strain OCU464* (NZ_CP009360.4) genomes using a custom python script.

### Statistical analysis and data availability

All statistical analyses were performed in R 3.5.0. Boxes in the boxplots indicate the 25th and 75th percentiles, the band in the box represents the median, and whiskers extend to 1.5 x interquartile range of the hinge. The sequence data for the non-endemic urine cohort was deposited in the database of Genotypes and Phenotypes (dbGaP, accession number phs001564v3.p1). The sequence data for the non-endemic plasma cohort was deposited in the Sequence Read Archive (accession number PRJNA263522). The sequence data generated in the scope of this study will be deposited in the Sequence Read Archive.

## Data Availability

The sequence data for the non-endemic urine cohort was deposited in the database of Genotypes and Phenotypes (dbGaP, accession number phs001564v3.p1). The sequence data for the non-endemic plasma cohort was deposited in the Sequence Read Archive (accession number PRJNA263522). The sequence data generated in the scope of this study will be deposited in the Sequence Read Archive.

## Acknowledgments

This work was supported by NIH Awards 1DP2AI138242 (to I.D.V.), R01AI146165 (to I.D.V.), 1R01AI151059 (to I.D.V.), a Synergy award from the Rainin Foundation (to I.D.V.), and a grant from the Bill and Melinda Gates Foundation INV-003145 (to I.D.V.). A.C. was supported by the National Institutes of Health under the Ruth L. Kirschstein National Research Service Award (6T32GM008267) from the National Institute of General Medical Sciences. P.B. was supported by the National Science Foundation under the Graduate Research Fellowships Program (DGE-1144153). A special thanks to the late Dr. Anna Lena Lopez for her research supervision with the Institute of Child Health and Human Development at the University of the Philippines Manila’s National Institutes of Health for samples collected and characterized in Manila, Philippines.

## Author’s Contributions

A. Chang, A.S., and I.D.V. contributed to the study design. A. Chang, O.M., L.D.K., J.L., and P.B. performed the experiments. A Chang, P.K., and I.D.V. analyzed the data. A.A., A. Cattamanchi, C.M.B., and J.C. facilitated data collection. A. Chang and I.D.V. wrote the manuscript. All authors provided comments and edits.

