## Supplemental Information for "A needle in a haystack: metagenomic DNA sequencing to quantify *Mycobacterium tuberculosis* DNA and diagnose tuberculosis"

**Table S1.** Description of databases tested for metagenomic classification. The genome assemblies are provided in Supplementary Data 1.

| **Name** | **Description** |
| --- | --- |
| MYCO2015 | All *Mycobacterium* complete genomes submitted after 2015 |
| MYCO_REF | All *Mycobacterium* reference genomes |
| TB2015 | All *M. tuberculosis* complete genomes submitted after 2015 |
| TB_REF | *M. tuberculosis* *H37Rv* reference genome |
| NTM_PD | Pulmonary disease-causing *Mycobacterium* reference genomes |
| NTM_ALL | Disease-causing *Mycobacterium* reference genomes |
| BAC_MYCO | All *Mycobacterium* reference genomes identified by RefSeq and the Pathosystems Resource Integration Center (PATRIC) pathogenic reference bacterial genomes |
| BAC_TB | *M. tuberculosis H37Rv* reference genome and the PATRIC pathogenic reference bacterial genomes |
| BAC_NTM | Non-tuberculous *Mycobacterium* and the PATRIC pathogenic reference bacterial genomes |
| BAC_PD | Pulmonary disease-causing *Mycobacterium* reference genomes and the PATRIC pathogenic reference bacterial genomes |

**Table S2.** Diagnostic performance summary of the 10 different databases with and without removal of confounding ribosomal RNA sequences for urine and plasma given as area under the curve values.

| **Database** | **Plasma**  **Sputum positive vs Sputum negative** | | **Plasma**  **Sputum positive vs Non-endemic** | | **Urine**  **Sputum positive vs Sputum negative** | | **Urine**  **Sputum positive vs Non-Endemic** | |
| --- | --- | --- | --- | --- | --- | --- | --- | --- |
|  | **All** | **-rRNA** | **All** | **-rRNA** | **All** | **-rRNA** | **All** | **-rRNA** |
| **TB2015** | 0.59 | 0.6 | 0.82 | 0.79 | 0.62 | 0.62 | 0.81 | 0.82 |
| **TB_REF** | 0.72 | 0.7 | 1 | 0.96 | 0.65 | 0.63 | 0.84 | 0.91 |
| **NTM_PD** | 0.72 | 0.68 | 1 | 0.96 | 0.65 | 0.65 | 0.85 | 0.91 |
| **NTM_ALL** | 0.72 | 0.69 | 1 | 0.97 | 0.63 | 0.62 | 0.84 | 0.91 |
| **MYCO2015** | 0.62 | 0.62 | 0.3 | 0.33 | 0.61 | 0.6 | 0.8 | 0.79 |
| **MYCO_REF** | 0.63 | 0.61 | 0.97 | 0.97 | 0.61 | 0.6 | 0.91 | 0.93 |
| **BAC_TB** | 0.71 | 0.69 | 1 | 0.96 | 0.62 | 0.61 | 0.91 | 0.93 |
| **BAC_PD** | 0.7 | 0.67 | 1 | 0.96 | 0.62 | 0.62 | 0.91 | 0.92 |
| **BAC_NTM** | 0.7 | 0.68 | 0.99 | 0.97 | 0.62 | 0.61 | 0.91 | 0.93 |
| **BAC_MYCO** | 0.62 | 0.6 | 0.97 | 0.97 | 0.6 | 0.59 | 0.94 | 0.94 |

**Table S3.** Overview of diagnostic performance of published nucleic acid assay for tuberculosis diagnostics in plasma and urine (NR = not reported).

| **Author** | **Year** | **Biofluid** | **Target** | **Amplicon Length** | **Sensitivity** | **Specificity** | **Location** | **Cohort** |
| --- | --- | --- | --- | --- | --- | --- | --- | --- |
| Patel^1^ | 2018 | Urine | DR region | 38 bp | 42.90% | 88.60% | South Africa | 175 Culture Positive |
|  |  |  |  |  |  |  |  | 238 Culture Negative |
| Ushio^2^ | 2016 | Plasma | IS6110 | 71 bp | 65% | 93% | Japan | 24 Culture Positive |
|  |  |  |  |  |  |  |  | 15 Healthy |
| Ushio^2^ | 2016 | Plasma | gyrB | 137 bp | 29% | 100% | Japan | 24 Culture Positive |
|  |  |  |  |  |  |  |  | 15 Healthy |
| Click^3^ | 2018 | Plasma | IS6110 | 106 bp | 45% | 67% | Kenya | 47 Culture Positive |
|  |  |  |  |  |  |  |  | 3 Culture Negative |
| Cannas^4^ | 2008 | Urine | IS6110 | 67 bp | 79% | 100% | Italy | 43 Culture Positive |
|  |  |  |  |  |  |  |  | 10 Culture Negative |
|  |  |  |  |  |  |  |  | 13 Healthy |
| Fortun^5^ | 2014 | Urine | 16S-rRNA | NR | 17.90% | NR | Spain | 28 Culture Positive |
| Labugger^6^ | 2017 | Urine | IS6110 | 38 bp | 64% | 100% | Germany | 11 Culture Positive |
|  |  |  |  |  |  |  |  | 8 Culture Negative |
| Torrea^7^ | 2006 | Urine | IS6110 | NR | 40.60% | 98.20% | Burkina Faso | 210 Culture Positive |
|  |  |  |  |  |  |  |  | 55 Culture Negative |
| Rebollo^8^ | 2006 | Urine | IS6110 | 123 bp | 42% | 100% | Spain | 43 Culture Positive |
|  |  |  |  |  |  |  |  | 14 Culture Negative |
|  |  |  |  |  |  |  |  | 13 Healthy |
|  |  |  |  |  |  |  |  | 13 Other Disease |
| Kafwabulula^9^ | 2002 | Urine | NR | NR | 55.60% | 98.40% | Zambia | 63 Culture Positive |
|  |  |  |  |  |  |  |  | 63 Culture Negative |
| Oreskovic^10^ | 2020 | Urine | IS6110 | 40 bp | 83.70% | 100% | South Africa | 49 Culture Positive |
|  |  |  |  |  |  |  |  | 10 Culture Negative |
|  |  |  |  |  |  |  |  | 14 Healthy Non-Endemic |

**Table S4.** Classification of synthetic *M. tuberculosis* reads at varying read coverages in urine samples from non-endemic (n=20, 3 replicates per sample) and tuberculosis (n=20, 3 replicates per sample) cohorts after removal of either host or host and *Mycobacterium* reads.

| **Cohort** | **Data** | **N** | **# Simulated** | **# Mapped** | **Accuracy** | **Precision** | **Sensitivity** | **F1** |
| --- | --- | --- | --- | --- | --- | --- | --- | --- |
| Non-endemic | Non-host | 60 | 2.69 | 2.69 | 0.977 | 0.844 | 0.977 | 0.889 |
| Non-endemic | Non-host | 60 | 5.86 | 5.78 | 0.981 | 0.912 | 0.987 | 0.939 |
| Non-endemic | Non-host | 60 | 13.6 | 13.4 | 0.98 | 0.953 | 0.988 | 0.968 |
| Non-endemic | Non-host | 60 | 158 | 156 | 0.974 | 0.975 | 0.992 | 0.982 |
| Non-endemic | Non-host, Non-*Mycobacterium* | 60 | 2.69 | 2.67 | 0.95 | 0.966 | 0.966 | 0.966 |
| Non-endemic | Non-host, Non-*Mycobacterium* | 60 | 5.86 | 5.83 | 0.978 | 1 | 0.994 | 0.997 |
| Non-endemic | Non-host, Non-*Mycobacterium* | 60 | 13.6 | 13.5 | 0.977 | 1 | 0.993 | 0.995 |
| Non-endemic | Non-host, Non-*Mycobacterium* | 60 | 158 | 156 | 0.973 | 1 | 0.995 | 0.997 |
| Tuberculosis | Non-host | 60 | 16.9 | 16.7 | 0.972 | 0.216 | 0.988 | 0.309 |
| Tuberculosis | Non-host | 60 | 54.5 | 53.9 | 0.972 | 0.375 | 0.986 | 0.491 |
| Tuberculosis | Non-host | 60 | 173 | 171 | 0.975 | 0.799 | 0.989 | 0.857 |
| Tuberculosis | Non-host | 60 | 888 | 878 | 0.975 | 0.799 | 0.989 | 0.857 |
| Tuberculosis | Non-host, Non-*Mycobacterium* | 60 | 16.9 | 16. | 0.974 | 0.995 | 0.984 | 0.989 |
| Tuberculosis | Non-host, Non-*Mycobacterium* | 60 | 54.5 | 53.9 | 0.973 | 1 | 0.988 | 0.994 |
| Tuberculosis | Non-host, Non-*Mycobacterium* | 60 | 173 | 171 | 0.977 | 1 | 0.986 | 0.993 |
| Tuberculosis | Non-host, Non-*Mycobacterium* | 60 | 888 | 878 | 0.976 | 1 | 0.989 | 0.994 |

**Table S5.** Classification of synthetic *M. bovis* reads at varying read coverages in urine samples from non-endemic (n=20, 3 replicates per sample) and tuberculosis (n=20, 3 replicates per sample) cohorts after removal of either host or host and *Mycobacterium* reads.

| **Cohort** | **Data** | **N** | **# Simulated** | **# Mapped** | **Accuracy** | **Precision** | **Sensitivity** | **F1** |
| --- | --- | --- | --- | --- | --- | --- | --- | --- |
| Non-endemic | Non-host | 60 | 2.69 | 2.69 | 0.989 | 0.827 | 0.98 | 0.88 |
| Non-endemic | Non-host | 60 | 5.86 | 5.78 | 1 | 0.903 | 0.99 | 0.938 |
| Non-endemic | Non-host | 60 | 13.6 | 13.4 | 1 | 0.951 | 0.987 | 0.967 |
| Non-endemic | Non-host | 60 | 158 | 156 | 1 | 0.976 | 0.979 | 0.975 |
| Non-endemic | Non-host, Non-*Mycobacterium* | 60 | 2.69 | 2.67 | 1 | 0.948 | 1 | 0.97 |
| Non-endemic | Non-host, Non-*Mycobacterium* | 60 | 5.86 | 5.83 | 0.997 | 0.974 | 0.981 | 0.974 |
| Non-endemic | Non-host, Non-*Mycobacterium* | 60 | 13.6 | 13.5 | 0.999 | 0.989 | 0.99 | 0.989 |
| Non-endemic | Non-host, Non-*Mycobacterium* | 60 | 158 | 156 | 0.999 | 0.995 | 0.992 | 0.989 |
| Tuberculosis | Non-host | 60 | 16.9 | 16.7 | 1 | 0.199 | 0.985 | 0.28 |
| Tuberculosis | Non-host | 60 | 54.5 | 53.9 | 1 | 0.359 | 0.989 | 0.478 |
| Tuberculosis | Non-host | 60 | 173 | 171 | 1 | 0.566 | 0.988 | 0.675 |
| Tuberculosis | Non-host | 60 | 888 | 878 | 1 | 0.796 | 0.989 | 0.856 |
| Tuberculosis | Non-host, Non-*Mycobacterium* | 60 | 16.9 | 16. | 0.999 | 1 | 0.993 | 0.996 |
| Tuberculosis | Non-host, Non-*Mycobacterium* | 60 | 54.5 | 53.9 | 1 | 1 | 0.991 | 0.995 |
| Tuberculosis | Non-host, Non-*Mycobacterium* | 60 | 173 | 171 | 1 | 1 | 0.988 | 0.994 |
| Tuberculosis | Non-host, Non-*Mycobacterium* | 60 | 888 | 878 | 1 | 1 | 0.989 | 0.994 |

**Table S6.** Classification of synthetic *M. avium* reads at varying read coverages in urine samples from non-endemic (n=20, 3 replicates per sample) and tuberculosis (n=20, 3 replicates per sample) cohorts after removal of either host or host and *Mycobacterium* reads.

| **Cohort** | **Data** | **N** | **# Simulated** | **# Mapped** | **Accuracy** | **Precision** | **Sensitivity** | **F1** |
| --- | --- | --- | --- | --- | --- | --- | --- | --- |
| Non-endemic | Non-host | 60 | 2.69 | 2.69 | 0.971 | 0.629 | 0.966 | 0.713 |
| Non-endemic | Non-host | 60 | 5.86 | 5.78 | 0.98 | 0.745 | 0.968 | 0.818 |
| Non-endemic | Non-host | 60 | 13.6 | 13.4 | 0.976 | 0.846 | 0.941 | 0.882 |
| Non-endemic | Non-host | 60 | 158 | 156 | 0.983 | 0.924 | 0.934 | 0.922 |
| Non-endemic | Non-host, Non-*Mycobacterium* | 60 | 2.69 | 2.67 | 0.95 | 0.761 | 0.929 | 0.823 |
| Non-endemic | Non-host, Non-*Mycobacterium* | 60 | 5.86 | 5.83 | 0.947 | 0.852 | 0.912 | 0.869 |
| Non-endemic | Non-host, Non-*Mycobacterium* | 60 | 13.6 | 13.5 | 0.973 | 0.94 | 0.946 | 0.937 |
| Non-endemic | Non-host, Non-*Mycobacterium* | 60 | 158 | 156 | 0.98 | 0.877 | 0.947 | 0.908 |
| Tuberculosis | Non-host | 60 | 16.9 | 16.7 | 0.981 | 0.197 | 0.946 | 0.292 |
| Tuberculosis | Non-host | 60 | 54.5 | 53.9 | 0.979 | 0.366 | 0.951 | 0.503 |
| Tuberculosis | Non-host | 60 | 173 | 171 | 0.981 | 0.621 | 0.946 | 0.74 |
| Tuberculosis | Non-host | 60 | 888 | 878 | 0.98 | 0.877 | 0.947 | 0.908 |
| Tuberculosis | Non-host, Non-*Mycobacterium* | 60 | 16.9 | 16. | 0.973 | 0.99 | 0.928 | 0.954 |
| Tuberculosis | Non-host, Non-*Mycobacterium* | 60 | 54.5 | 53.9 | 0.977 | 1 | 0.943 | 0.969 |
| Tuberculosis | Non-host, Non-*Mycobacterium* | 60 | 173 | 171 | 0.981 | 1 | 0.946 | 0.972 |
| Tuberculosis | Non-host, Non-*Mycobacterium* | 60 | 888 | 878 | 0.981 | 1 | 0.946 | 0.972 |

**Table S7.** Insertion sequences used to classify the nontuberculous mycobacterial background.

| **Species** | **Insertion sequences** | **Accession** |
| --- | --- | --- |
| *Mycobacterium avium* | IS1110 | Z23003.1 |
|  | IS1245 | L33879 |
|  | IS1311 | CP000479 |
|  | IS1601 | CP000479 |
|  | IS1612 | AJ251813 |
|  | IS902 | X58030 |
|  | IS666 | AF107207 |
|  | IS999 | AF232829 |
| *Mycobacterium branderi* | IS1408 | U62766 |
| *Mycobacterium celatum* | IS1407 | X97307 |
| *Mycobacterium fortuitum* | IS219 | MF018875 |
|  | IS220 | AJ315500 |
| *Mycobacterium gordonae* | IS1511 | U95315 |
|  | IS1512 | U95314 |
| *Mycobacterium intracellulare* | IS1141 | L10239 |
| *Mycobacterium smegmatis* | IS1096 | M76495 |
|  | IS1137 | X70913 |
|  | IS1549 | CP000480 |
|  | IS6120 | M69182 |
| *Mycobacterium ulcerans* | IS2404 | AF003002 |
| *Mycobacterium xenopi* | IS1395 | U35051 |

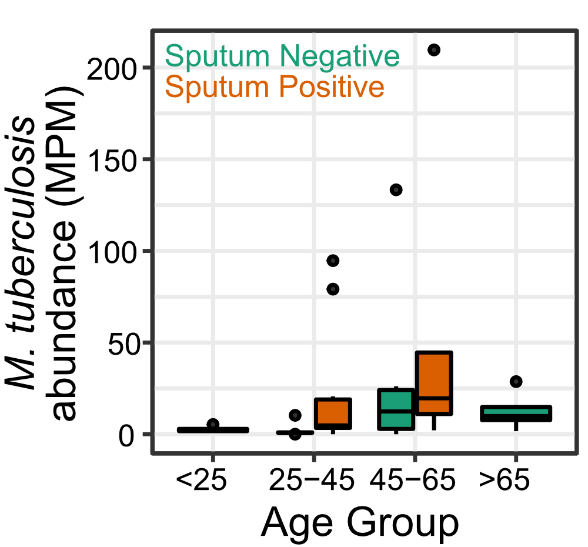

**Figure S1.** Correlations between age and *M. tuberculosis* abundance.

**Description of Additional Supplementary Files**

File Name: Supplementary Data 1 (myco_databases.xslx)

Description: Genome assemblies used to create the different databases used for metagenomic classification.

10.	Oreskovic, A. *et al.* Diagnosing Pulmonary Tuberculosis by Using Sequence-Specific Purification of Urine Cell-Free DNA. *J. Clin. Microbiol.* **59**, e00074-21.
